# Virological characteristics of SARS-CoV-2 vaccine breakthrough infections in health care workers

**DOI:** 10.1101/2021.08.20.21262158

**Authors:** Marc C. Shamier, Alma Tostmann, Susanne Bogers, Janet de Wilde, Jeroen IJpelaar, Willemijn A. van der Kleij, Herbert de Jager, Bart L. Haagmans, Richard Molenkamp, Bas. B. Oude Munnink, Carsten van Rossum, Janette Rahamat-Langendoen, Nannet van der Geest, Chantal P. Bleeker-Rovers, Heiman Wertheim, Marion P.G. Koopmans, Corine H. GeurtsvanKessel

**Affiliations:** Department of Viroscience, Erasmus Medical Center, Rotterdam, The Netherlands; Department of Medical Microbiology, Radboud Centre for Infectious Diseases, Radboud university medical center, Nijmegen, The Netherlands; Department of Occupational Health Services, Erasmus Medical Center, Rotterdam, Netherlands; Department of Internal Medicine, Radboud Centre for Infectious Diseases, Radboud university medical center, Nijmegen, The Netherlands; Department of Occupational Health, Radboud university medical center, Nijmegen, The Netherlands

## Abstract

**Background:** SARS-CoV-2 vaccines are highly effective at preventing COVID-19-related morbidity and mortality. As no vaccine is 100% effective, breakthrough infections are expected to occur.

**Methods:** We analyzed the virological characteristics of 161 vaccine breakthrough infections in a population of 24,706 vaccinated healthcare workers (HCWs), using RT-PCR and virus culture.

**Results:** The delta variant (B.1.617.2) was identified in the majority of cases. Despite similar Ct-values, we demonstrate lower probability of infectious virus detection in respiratory samples of vaccinated HCWs with breakthrough infections compared to unvaccinated HCWs with primary SARS-CoV-2 infections. Nevertheless, infectious virus was found in 68.6% of breakthrough infections and Ct-values decreased throughout the first 3 days of illness.

**Conclusions:** We conclude that rare vaccine breakthrough infections occur, but infectious virus shedding is reduced in these cases.

## Introduction

Once COVID-19 vaccines became available, health care workers (HCWs) were among the first groups to be vaccinated and reach high vaccine coverage. Registered vaccines have been highly effective in preventing clinically significant coronavirus disease 2019 (COVID-19), caused by severe acute respiratory syndrome coronavirus 2 (SARS-CoV-2)^1^, and have shown to reduce the incidence of infections^2,3^. However, mild breakthrough infections in a small percentage of vaccine recipients have been described^4-9^ which is not surprising as none of the registered vaccines will provide sterile immunity against infection^10-13^. In a setting of mass vaccination, the BNT162b2 vaccine was highly effective (92%) at preventing infection from 7 days after the second dose^14^, but a recent study from Israel described vaccine breakthrough infections in 39 health care workers vaccinated with the BNT162b2 mRNA vaccine. The alpha variant was identified as the main causative strain and a majority of cases presented low Ct-values (<30), indicating probable infectivity^4^. For the single dose Ad26.COV2.S adenoviral vector vaccine, a phase IV study reported a 76.1% effectiveness to prevent infection from 14 days after vaccination^15^. The effectiveness against infection with the delta (B.1.617.2) variant was 88% for the BNT162B2 vaccine and 67% for the ChAdOx1 vaccine, moderately lower than against infection with the alpha (B.1.1.7) variant^16^.

Up to present, little is known about the virological kinetics of SARS-CoV-2 breakthrough infections, and the role of the vaccinated host in the transmission cycle. Better understanding of the dynamics of breakthrough infections is essential to define infection prevention and (public) health policies during the next phase of the pandemic. In this study, we report the virological findings of 161 vaccine breakthrough infections occurring from April to July 2021 in the Netherlands. The infections occurred in HCWs working in two tertiary care hospitals, who were immunized with various mRNA and viral vector vaccines. Infections were caused predominantly by the SARS-CoV-2 delta variant and virus culture was performed as a proxy for infectivity.

## Results

A total of 161 fully vaccinated HCWs diagnosed with COVID-19 by PCR were included in this study. In accordance with case definitions defined by the Centers for Disease Control and Prevention, infections were classified as breakthrough infections if the date of the first positive SARS-CoV-2 RT-PCR was more than 14 days after completion of all recommended vaccine doses^17^. Cases with symptom onset <14 days after the last vaccine dose and cases with a previous positive test <45 days prior were not considered breakthrough infections. In parallel with a surge in cases in the general Dutch population^18^, an increased incidence of breakthrough infections in HCWs was observed in July 2021. In 126 samples a SARS-CoV-2 lineage could be identified, 90.5% of these showed presence of the delta variant. The mean age of the HCWs with a breakthrough infection was 25.5 years and 91% were less than 50 years old (Table 1). All infections were mild and did not require hospital admission. The individuals were vaccinated between January and May 2021 with either an mRNA vaccine or a viral vector vaccine. Table 1 shows the distribution of vaccines among all HCWs and among the HCWs with breakthrough infections. Although the data may imply an overrepresentation of Ad26.COV2.S and BNT162b2 vaccine recipients among the cases, this study was not designed to compare vaccine effectivity. The indication to receive a certain vaccine was not random and data on risk factors for exposure were not recorded, therefore potential confounders could not be adjusted for.

**Table 1.**
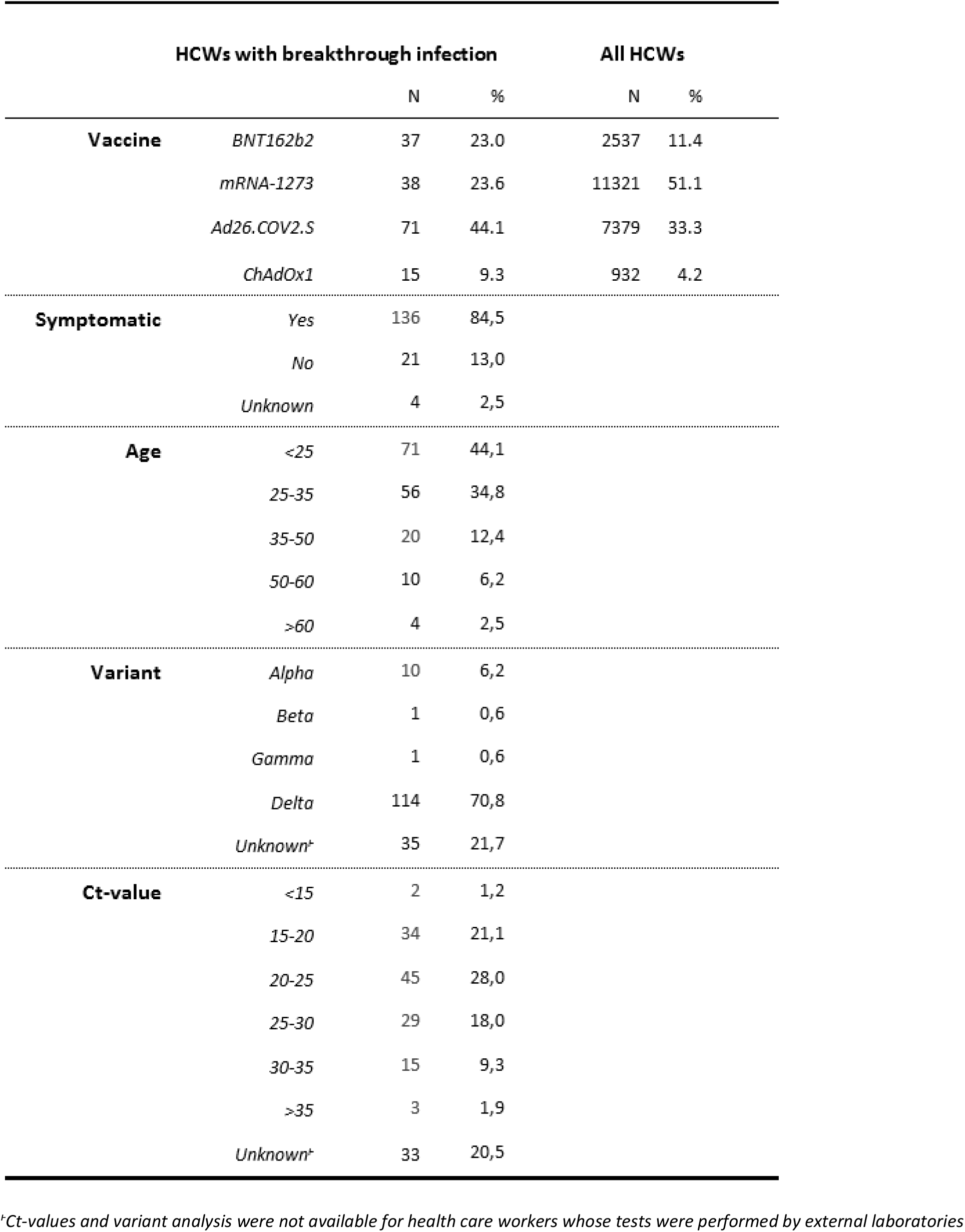
Characteristics of health care workers with SARS-CoV-2 vaccine breakthrough infections.

Table 1 shows the distribution of Ct-values of the breakthrough infections, as a proxy for the nasopharyngeal viral load. Ct-values were significantly lower in symptomatic breakthrough infections (μ = 23.2) than in asymptomatic breakthrough infections (μ = 26.7), corresponding to higher viral loads (p = 0.022, t-test). In symptomatic vaccinated HCWs, the Ct-values decreased significantly throughout the first days from symptom onset and were lowest on the third day of illness (Figure 1A). There were no statistically significant differences in Ct-values between HCWs immunized with the 4 different vaccines. Furthermore, the time since the administration of the last vaccine dose showed no clear relationship with Ct-values (R^2^ = 0.02, p = 0.13, linear regression).

**Figure 1.**
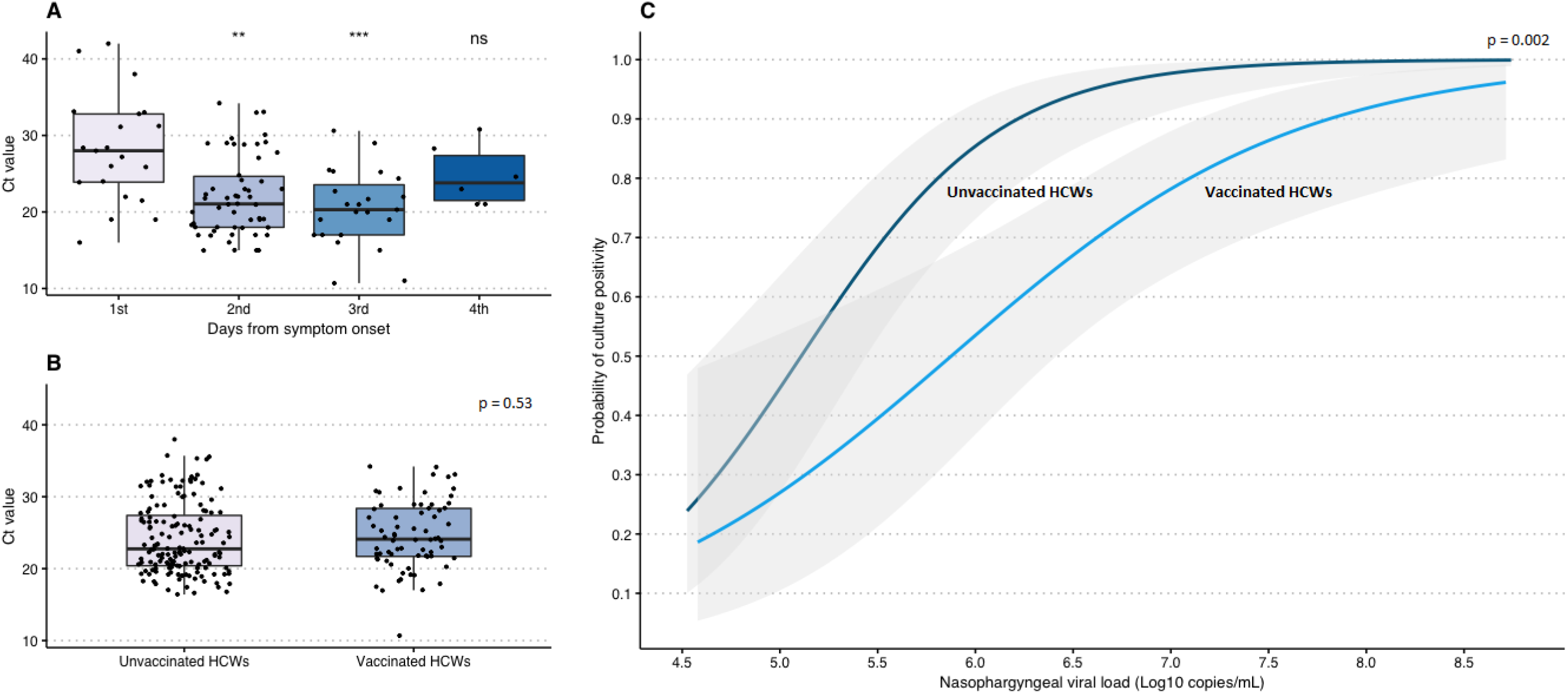
SARS-CoV-2 culture positivity and Ct-values in nasopharyngeal samples of health care workers with SARS-CoV-2 breakthrough infections. (A) Ct-values by day from symptom onset (B) Ct-values of HCWs with vaccine breakthrough infections (primarily Delta) compared to Ct-values of HCWs with primary infections (primarily D614G) (C) Probability of culture positivity by nasopharyngeal viral load (Probit Analysis), comparing HCWs with vaccine breakthrough infections (primarily Delta) to HCWs with primary infections (primarily D614G)

Subsequently, RT-PCR positive swabs were tested for the presence of infectious virus using cell culture. As a reference, we used the (first positive) samples from mild primary infections that occurred in the same cohort of HCWs prior to the onset of vaccination, these infections were primarily caused by SARS-CoV-2 D614G. The mean Ct-value upon diagnosis was similar between these two groups: 24.6 (15.3 - 33.9) for vaccinated HCWs and 24.2 (14.53 - 33.8) for unvaccinated HCWs (p = 0.53, t-test) (Figure 1B). The SARS-CoV-2 culture of nasopharyngeal swabs was positive in 68.6% of vaccinated HCWs versus 84.9% of unvaccinated HCWs with primary infections (p = 0.005, t-test). As the probability of culture positivity depends on viral load^19^, this was corrected for using a probit regression model with both viral load and vaccination status as predictors. Figure 1C shows the probability of a positive culture for a given viral load in vaccinated and unvaccinated HCWs. A positive vaccination status significantly decreased the probability of culture positivity (p = 0.002, Wald test).

## Discussion

In this study we assessed the virological kinetics of mild COVID-19 breakthrough infections upon immunization with several vaccines. Our data support that the SARS-CoV-2 infectious virus shedding is lower in vaccinated individuals with breakthrough infections (caused by primarily the delta variant) than in unvaccinated individuals with primary infections (caused by SARS-CoV-2 D614G). Nevertheless, virus culture was positive in 68.6% of breakthrough infections and Ct-values decreased throughout the first three days of illness. Despite the reduced viral viability, the infectivity of individuals with breakthrough infections should not be neglected.

It remains a challenge to assess infectivity of an individual based on clinical sampling. Although RT-PCR is a highly sensitive method to diagnose SARS-CoV-2 infection, it detects RNA produced in infected cells rather than whole (infectious) virions and is therefore not an optimal indicator of infectivity ^19^. Ct-values are sometimes used to differentiate between phases of infection, but the definition of a cut-off value is complicated, due to the large variety of assays and clinical samples. Considering these limitations of RT-PCR, demonstrating viral viability through replication in cell culture is currently considered the best proxy to demonstrate infectious virus in a clinical specimen^20,21^. We and others previously showed that the viability of SARS-CoV-2 depends on several factors among which the severity of disease, timing of sampling, the type of specimen and presence of antibodies^19,22^. To our knowledge this is the first study to report on virus cultures in COVID-19 vaccine breakthrough infections. Although reduced immune responses may likely account for breakthrough infections, further studies are needed to investigate whether these are still able to reduce infectious virus shedding.

Obviously, the use of virus culture has its limitations as well: it is a laborious method only performed in specialized BSL-3 laboratories and therefore not widely applicable. In addition, lack of standardization of methods (e.g. the cell line used) still hampers interchangeability of results between laboratories. Nevertheless, an experimental animal study on SARS-CoV-2 transmission recently confirmed a strong correlation between transmission and virus culture ^23^.

To study the effect of vaccination on infectivity, it would be preferable to compare infections occurring in vaccinated and unvaccinated individuals during the same time period, to minimize the impact of different SARS-CoV-2 variants. Due to the high vaccine coverage in HCW, we diagnosed very few infections in unvaccinated HCWs. For this reason, we used infections prior to the onset of vaccination as a reference. Although the predominant SARS-CoV-2 variant differed between groups, the groups were similar with respect to demographic characteristics, severity of disease, testing algorithms and Ct-values upon diagnosis. The study participants comprise a population immunized with several vaccines, which reflects the current situation in many countries. This study was not designed to detect differences in vaccine effectiveness, as HCWs who received different vaccines also differed with respect to demographic characteristics. The frequency of breakthrough infections in the different groups was likely influenced by variables that were not controlled for.

Phase IV studies have confirmed that vaccination is highly effective at preventing COVID-19-related morbidity and mortality^2,14,15^ although vaccine effectiveness will never reach 100%. Our study supports the excellent effectiveness of vaccination in preventing severe SARS CoV-2 related disease, but also demonstrates that vaccinated individuals can still acquire infection and carry infectious virus. Although symptomatic vaccinated individuals should be tested to further reduce the chance of virus transmission to individuals at risk for severe disease, further studies are needed to assess whether the decreased infectious virus shedding in breakthrough infections also lowers the chance of virus transmission.

## Methods

### Study population

Data were collected and analyzed anonymously from HCWs of two tertiary care centers in the Netherlands (Erasmus University Medical Center, Rotterdam and Radboud University Medical Center, Nijmegen), together employing over 25,000 HCWs. Since April 2020, approximately 1900 symptomatic HCWs presenting to the occupational health services department were enrolled into a prospective cohort study^24^. Symptomatic HCWs underwent questioning and SARS-CoV-2 RT-PCR testing, complemented by tracing and testing of contacts, resulting in detection of asymptomatic cases. Any infections diagnosed by external laboratories were reported by the department of the respective employee. In both centers, immunization of HCWs commenced in January 2021 with the BNT162b2 mRNA vaccine (Pfizer-BioNTech), prioritizing physicians and nurses working directly with COVID-19 cases. The majority of HCWs received either the mRNA-1273 (Moderna) or Ad26.COV2.S (Janssen) vaccines and a minority was vaccinated with ChAdOx1 (AstraZeneca Oxford) (Table 1). The indication of the different vaccines was based on availability and professional role. Throughout the study period, all HCWs with symptomatic breakthrough infections followed institutional infection prevention guidelines and resumed their professional activities only after full recovery. HCWs with asymptomatic breakthrough infections remained in home isolation for at least 3 days. We compared virological characteristics of first RT-PCR positive samples collected from HCWs with breakthrough infections to first RT-PCR positive samples from the same cohort of HCWs prior to the onset of vaccination. The breakthrough infections occurred between April and July 2021, the primary infections occurred between April and December 2020 and were primarily caused by SARS-CoV-2 D614G. The two groups did not differ with respect to demographic characteristics and testing algorithms remained unchanged.

### RT-PCR for the detection of SARS-CoV-2 RNA

SARS-CoV-2 RT-PCR tests were performed on nasopharyngeal swabs, using the SARS-CoV-2 test on a Cobas® 6800 system (Roche Diagnostics) in Erasmus MC and using the Aurora Flow (Roche Diagnostics) in Radboud Medical Center. Using a formula based on E-gene calibration curves, cycle threshold (Ct) values were converted to viral load in Log_10_ RNA copies/mL. This conversion method was previously validated^25^.

### Typing of SARS-CoV-2 variants using RT-PCR or next generation sequencing

All positive samples were analyzed for the presence of mutations indicative of variants of concern, either by next generation sequencing or by the use of variant-specific RT-PCR tests (VirSNiP assays 53-0799 and 53-0807, TIB Molbiol, Berlin, Germany) using a SYBR Green melting curve protocol. These assays screen for the presence of HV69/70 deletion, N501Y, E484K, K417T/K417N and P681R mutations. The results were interpreted based on the genomics of the SARS-CoV-2 lineages circulating at the time of this study. The combination of the HV69/70 deletion and N501Y mutation was considered indicative for the Alpha variant (B.1.1.7). A K417N/E484K/N501Y profile was considered indicative for the Beta variant (B.1.351), K417T/E484K/N501Y for the Gamma variant (P.1) and P681R for the Delta variant (B.1.617.2).

### Virus culture

Virus culture was performed on all samples collected in the Erasmus Medical Center, by inoculating Vero cells (clone 118) as previously described^19^. All cultures were performed in twofold, with one replicate for immunofluorescence analysis after acetone fixation at 48h of incubation. The second replicate was microscopically examined for the presence of cytopathic effect daily for 2 weeks. Viral culture was considered negative if no cytopathic effect was observed after 14 days of incubation. To investigate how the probability of the binary outcome (culture positivity) depends on viral load and vaccination, the culture results were analyzed using probit regression.

### Statistical analysis

All statistical analyses was performed using R Statistical Software version 4.1.1 (Foundation for Statistical Computing, Austria) and STATA statistical software program version 13.1 (Statacorp, USA).

### Ethical approval

This study was approved by Radboud university medical center Committee on Research Involving Human Subjects and the Erasmus Medical Center Medical Ethics Committee. All samples were collected following routine institutional COVID-19 testing guidelines, the participants were not subject to any procedures for the purpose of this study and all data were anonymized prior to analysis.

## Data Availability

Data are available upon request

## Acknowledgements

David van de Vijver, Jolanda Kreeft-Voermans, Amber Weevers, Anoushka Comvalius, Djenolan van Mourik and Michael van der Voorden are gratefully acknowledged for their technical and analytical contributions.

## Notes

### Competing Interest Statement

The authors have declared no competing interest.

### Funding Statement

No external funding

### Author Declarations

This study was approved by Radboud university medical center Committee on Research Involving Human Subjects (CMO) and the Erasmus Medical Center Medical Ethics Committee (METC). All samples were collected following routine institutional COVID-19 testing guidelines, the participants were not subject to any procedures for the purpose of this study and all data were anonymized prior to analysis.

